# Sex-specific associations between type 2 diabetes incidence and exposure to dioxin and dioxin-like pollutants: a meta-analysis

**DOI:** 10.1101/2021.09.28.21264274

**Authors:** Noa Gang, Kyle Van Allen, Paul J. Villeneuve, Heather MacDonald, Jennifer E. Bruin

**Author notes:** Address correspondence to: Dr. Jennifer Bruin, 1125 Colonel By Drive, Ottawa, ON K1S 5B6.

## Abstract

The relationship between persistent organic pollutants (POPs), including dioxins and dioxin-like polychlorinated biphenyls (DL-PCBs), and diabetes incidence in adults has been extensively studied. However, significant variability exists in the reported associations both between and within studies. Emerging data from rodent studies suggest that dioxin exposure disrupts glucose homeostasis in a sex-specific manner. Thus, we performed a meta-analysis of relevant epidemiological studies to investigate whether there are sex-specific associations between dioxin or DL-PCB exposure and type 2 diabetes incidence. Articles were organized into the following subcategories: data stratified by sex (16%), unstratified data (56%), and data from only 1 sex (16% male, 12% female). We also considered whether exposure occurred either abruptly at high levels through a contamination event (“disaster exposure”) or chronically at background levels (“non-disaster exposure”). Only 8 studies compared associations between dioxin/DL-PCB exposure and diabetes risk in males versus females within the same population. When all sex-stratified or single sex studies were considered in the meta-analysis, the summary odds ratio (OR) for increased diabetes risk was similar between females and males (1.78 and 1.95, respectively) when comparing exposed to reference populations, suggesting that this relationship is not sex-specific. However, when we considered disaster-exposed populations separately, the association differed substantially between sexes, with females showing a much higher OR than males (2.86 and 1.59, respectively). Moreover, the association between dioxin/DL-PCB exposure and diabetes was stronger for females than males in disaster-exposed populations. In contrast, both sexes had significantly increased ORs in non-disaster exposure populations and the OR for females was lower than males (1.40 and 2.02, respectively). Our review emphasizes the importance of considering sex differences, as well as the mode of pollutant exposure, when exploring the relationship between pollutant exposure and diabetes in epidemiological studies.

## 1. Introduction

Diabetes incidence is increasing worldwide at a rate that cannot be explained solely by genetic predisposition or lifestyle (Franks and McCarthy 2016; Butalia et al. 2016; Knip et al. 2005), prompting investigations into potential alternative sources of diabetes risk. There is emerging evidence pointing to an association between environmental pollutant exposure and type 2 diabetes (T2D) in humans (Carpenter 2008; Hectors et al. 2011; Ngwa et al. 2015; Taylor, David 2001). Persistent organic pollutants (POPs) are man-made toxins, released into the environment through industrial, electrical, and agricultural sources (Hens and Hens 2017; Wikoff, Fitzgerald, and Birnbaum 2012). POPs are typically lipophilic, resistant to degradation, and highly mobile, thus leading to ubiquitous global dispersion and bioaccumulation (Fisher 1999). Pollutant exposure can occur abruptly at high levels, as in a disaster event such as an industrial accident or food contamination, but more frequently occurs at chronic low levels (Marinković et al. 2010). Human exposure to POPs is typically through consumption of fish, meat, eggs, and dairy (Srogi 2008; Schecter et al. 2006). Despite global efforts to restrict POP production, use continues in some countries (Jaacks et al. 2019; Azandjeme et al. 2014) and biomonitoring studies continue to detect POPs in bodily fluids of the general population in North America (Haines et al. 2017; Haines and Murray 2012).

### 1.2 Emerging links between POPs and T2D pathogenesis

T2D is characterized by insufficient insulin production by pancreatic beta cells in the face of peripheral insulin resistance, which results in chronic hyperglycemia (Kahn 2003; Weir and Bonner-Weir 2004; Porte and Kahn E 2001; Chen et al. 2017; Kahn et al. 2009). This disease manifests slowly and early symptoms of metabolic dysfunction, such as impaired glucose tolerance or hyperinsulinemia, can last for years (Porte and Kahn E 2001; Weir and Bonner-Weir 2004; Chen et al. 2017). Clinical diagnoses of diabetes is internationally defined as glycated haemoglobin (HbA1c) ≥6.5%, fasting plasma glucose ≥7.0 mmol/L, or 2-hour plasma glucose during an oral glucose tolerance test (OGTT) of ≥11.1 mmol/L (Gillett 2009). Environmental factors that adversely impact beta cell health and/or peripheral insulin sensitivity could augment underlying vulnerabilities and promote the development of T2D.

Within the last decade numerous publications have reported positive associations between exposure to POPs and T2D incidence (A. Al-Othman et al. 2014; A. A. Al-Othman, Abd-Alrahman, and Al-Daghri 2015; Aminov, Haase, and Carpenter 2016; Aminov et al. 2016; Arrebola et al. 2013; Cappelletti et al. 2016; Eslami et al. 2016; Everett and Matheson 2010; Everett and Thompson 2012; Gasull et al. 2012; Han et al. 2020; Huang et al. 2015; D. H. Lee et al. 2010; 2011; Lind et al. 2014; Mannetje et al. 2018; Marushka et al. 2017; Patel, Bhattacharya, and Butte 2010; Rahman et al. 2019; Rylander et al. 2015; Singh and Chan 2017; Son et al. 2010; Starling et al. 2014; Suarez-Lopez et al. 2015; Tanaka et al. 2011; Tornevi et al. 2019; Ukropec et al. 2010; Wolf et al. 2019; Wu et al. 2013; Zong et al. 2016). However, the strength of associations between POPs and diabetes incidence varies considerably (D.-H. Lee et al. 2014; Hectors et al. 2011; Ngwa et al. 2015; C. Yang et al. 2017). There are numerous potential explanations for variability in the epidemiology literature, such as the type of POPs studied, exposure duration, level of exposure, method of contaminant analysis, underlying health and genetics of the exposed populations, ongoing medical treatments, how diabetes incidence is determined, and the range of covariates considered. In this meta-analysis, we narrowed down the scope of literature on environmental contaminants by focusing on studies that measured dioxins and dioxin-like (DL) POPs.

### 1.3 Dioxins and dioxin-like pollutants

Various chemicals are classified as POPs, including polychlorinated dibenzo-*p*-dioxins and dibenzofurans (PCDD/Fs), polychlorinated biphenyls (PCBs), organochlorine pesticides (OCPs), poly fluorinated alkyl substances (PFAS), and polybrominated diphenyl ethers (PBDEs). Dioxins and DL-PCBs are structurally similar polycyclic, halogenated aromatic chemicals (**Figure 1**) that share a common mechanism of action via binding to the intracellular aryl hydrocarbon receptor (AhR).

**Figure 1.**
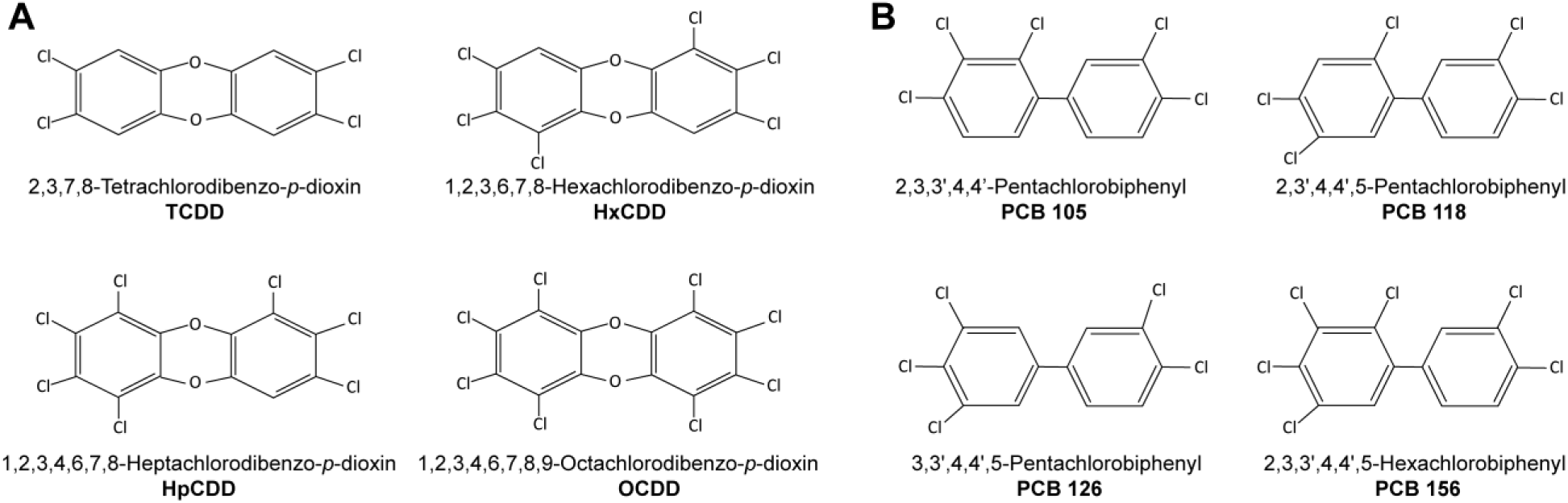
Chemical structure of (**A**) dioxins and (**B**) dioxin-like (DL) PCBs commonly considered in epidemiological studies.

The dioxin family consists of oxygen-linked chlorinated benzene rings that vary in position and number of halogens (**Figure 1A**). The toxicity of different dioxin compounds is related to their pattern of halogenation. For example, TCDD (2,3,7,8-tetrachlorodibenzo-*p-*dioxin; **Figure 1A**) is the most toxic and stable dioxin, with a half-life of 7-11 years in humans (D.-H. Lee et al. 2014; Schecter et al. 2006; Srogi 2008). During much of the 1900’s TCDD was produced as a contaminant from industrial processing, but was eventually recognized as a health hazard to industrial workers (Calvert et al. 1999; Cappelletti et al. 2016; Kyle Steenland et al. 1999; Vena et al. 1998) and local communities (Huang et al. 2015; Karnes, Winquist, and Steenland 2014; Chang et al. 2010; Ukropec et al. 2010; MacNeil et al. 2009; Karouna-Renier et al. 2007; Cranmer et al. 2000). Widespread distribution of TCDD occurred during the Vietnam war when Agent Orange, a defoliate contaminated with TCDD, was used by the US military as part of a chemical warfare program known as Operation Ranch Hand (Henriksen, Gary et al. 1997; Kim et al. 2003; Longnecker and Michalek 2000; Michalek and Pavuk 2008; K. Steenland et al. 2001). Furthermore, in 1976 an industrial disaster in Seveso, Italy released tonnes of nearly pure TCDD gas into the local regions. Other dioxins commonly considered within the literature and included in our review are 1,2,3,6,7,8-Hexachlorodibenzo-*p*-dioxin (HxCDD), 1,2,3,4,6,7,8-Heptachlorodibenzo-*p*-dioxin (HpCDD), and 1,2,3,4,6,7,8,9-Octachlorodibenzo-*p*-dioxin (OCDD) (**Figure 1A**). However, TCDD has been more widely studied for sub-lethal exposure effects compared to other dioxins, as it was involved in more disaster and occupational exposure events (Birnbaum and Couture 1988; Couture, Elwell, and Birnbaum 1988; Rozman 1999; Schwetz et al. 1973; Viluksela et al. 1997; 1998).

The PCB family includes 209 separate congeners, 12 of which are considered DL-PCBs (US Environmental Protection Agency 2003) (**Figure 1B**). PCBs are exceptionally stable, heat resistant and non-flammable, and were intentionally manufactured between the 1920’s to 70’s for electrical components, hydraulic fluids, lubricants, and industrial insulating or heat-exchange fluids (White and Birnbaum 2009; Wikoff, Fitzgerald, and Birnbaum 2012). Disaster exposure to DL-PCBs occurred in Michigan, US in 1973 when animal feed accidently contaminated with PCBs was distributed to farms (Vasiliu et al. 2006). Additionally, between 1978 to 1979, rice-bran oil contaminated with PCBs and PCDFs poisoned thousands of inhabitants of Yucheng, Taiwan with “oil disease” (S. L. Wang et al. 2008). These incidents of disaster exposure to DL-PCBs, albeit far from isolated events, are the most widely studied when considering pathologies related to metabolic diseases (Hens and Hens 2017; Taylor, David 2001).

### 1.4 Evidence for a causal link between dioxin exposure and diabetes

Dioxins and DL-PCBs act as ligands for AhR, leading to upregulation of target genes such as cytochrome P450 *(Cyp)1a1*. Our lab showed that TCDD-exposed mice have persistent CYP1A1 upregulation in pancreatic islets, where insulin-secreting beta cells reside, a sign that TCDD reaches the endocrine pancreas *in vivo* (Ibrahim et al. 2020). A single high-dose injection of TCDD in mice caused hypoinsulinemia *in vivo* for up to 6 weeks and reduced glucose-stimulated insulin secretion in islets *ex vivo* (Hoyeck et al. 2020; Ibrahim et al. 2020). Furthermore, direct TCDD exposure *in vitro* caused *CYP1A1* upregulation and suppressed glucose-induced insulin secretion in both mouse and human islets (Ibrahim et al. 2020; Kurita et al. 2009). These data suggest that TCDD may be driving metabolic dysfunction, at least in part, via direct effects on pancreatic islets.

There is also emerging evidence that female rodents are more susceptible to the diabetogenic effects of dioxins compared with male rodents (Matteo et al. 2020; Hoyeck et al. 2020; Naville et al. 2013). For example, while a single high-dose injection of TCDD at 8 weeks of age caused persistent hypoinsulinemia in both male and female mice *in vivo*, only male mice had increased insulin sensitivity and altered islet cell composition, and only female mice developed transient hyperglycemia following TCDD exposure (Hoyeck et al. 2020). Consistent with the single high-dose model, repeated low-dose TCDD exposure for 12 weeks starting at 6-8 weeks of age both exacerbated and accelerated the onset of high fat diet (HFD)-induced glucose intolerance in female but not male mice (Matteo et al. 2020). Naville et al. (2013) also observed exacerbated HFD-induced glucose intolerance in female, but not male mice, exposed chronically from 5-10 weeks of age to a low-dose mixture of pollutants that included TCDD.

Considering the sex differences reported in mouse models, we hypothesized that there may also be sex-specific associations between dioxin/DL pollutant exposure and T2D incidence in humans. To explore this question, we performed a meta-analysis of epidemiological studies that assessed exposure to either dioxins or DL-PCBs and incidence of T2D or metabolic syndrome. We also considered the mode of pollutant exposure, where a disaster event (e.g. Seveso) reflects abrupt, high-dose exposure versus a non-disaster chronic exposure scenario, ranging from low-dose background levels (e.g. general population) to higher-dose exposure (e.g. occupational settings). Importantly, abrupt exposure via disaster can lead to a prolonged period of high concentration pollutant exposure due to the long half-life of dioxins and DL pollutants (Fisher, 1999).

## 2. Methods

### 2.1 Literature search strategy

We conducted a literature search for studies examining POPs, dioxins, or benzofurans and diabetes in humans via PubMed on May 17, 2021. MeSH terms, substance registry numbers, and keywords were used. The search terms can be found in **Supplementary Table 1**. Review articles were excluded unless original data were presented. A filter for human studies was applied for studies published before 2020. A simple secondary search was conducted in Google Scholar to supplement the PubMed search.

Records were screened by two independent reviewers using the following inclusion criteria: the population included human adults exposed to dioxins, DL-PCBs, or benzofurans, and an outcome of T2D, hyperglycemia, prediabetes, metabolic syndrome, or glucose intolerance.

### 2.2 Data extraction

Data extracted from each study included: authors, publication year, cohort location, type of study (e.g. cohort, cross-sectional, case-control), sample size, type of exposure (disaster or non-disaster), specific pollutant(s) measured, diabetes assessment method, outcome determined, considerations made for sex-specific associations (whether data was stratified by sex, unstratified by sex, or only considered 1 sex), analysis strategy (e.g. covariates), and measures of association (see **Table 1** for key details and **Supplementary Table 2** for additional information on each study). Study quality was determined by 2 independent assessors. All studies included in the meta-analysis were assessed for publication bias via funnel plot (**Supplementary Figures 1-3**). Primary summary measures were usually stated as risk ratio (RR) or odds ratio (OR), however incidence rate ratio (IRR), and incidence density ratio (IDR) association measures were also included.

**Table 1.** Summary of 18 articles investigating the association between dioxins and/or DL-PCBs and diabetes incidence included in the meta-analysis.

|  | Reference | Study Participants | Outcome | Pollutant | Mode of exposure | Measure of Association (OR, RR, IDR, IRR) |
| --- | --- | --- | --- | --- | --- | --- |
| **Sex-stratified studies (8 articles)** | **Bertazzi et al. 2001** | Males (n=22,607)<br>Females (n= 22,762) | T2D | TCDD | Disaster (plant explosion) | RR (95% CI) = 0.6 (0.1-4.1) males<br>RR (95% CI) = 2.2 (1.0-4.6) females |
|  | **Han et al. 2020** | Males (n=151)<br>Females (n=165) | T2D | PCB-118 | Non-disaster (low background) | OR (95% CI) = 4.62 (1.52-14.06) males<br>OR (95% CI) = 3.08 (0.92-10.35) females |
|  | **Huang et al. 2015** | n=2,898 (total participants) | T2D | PCDD/Fs | Non-disaster (high background) | OR (95% CI) = 3.3 (2.0-5.5) males<br>OR (95% CI) = 2.3 (1.2-4.4) females |
|  | **Huang et al. 2017** | Males (n=1,453)<br>Females (n=1,305) | Metabolic Syndrome | PCDD/Fs | Non-disaster (high background) | OR (95% CI) = 1.59 (1.22-2.08) males<br>OR (95% CI) = 1.16 (0.8-1.68) females |
|  | **Silverstone et al. 2012** | Males (n=153)<br>Females (n=427) | T2D | DL-PCBs | Non-disaster (high background) | OR (95% CI) = 0.71 (0.36-1.39) males<br>OR (95% CI) = 1.51 (1.02-2.24) females |
|  | **Turyk et al. 2009** | Males (n=279)<br>Females (n=192) | T2D | PCB-118 | Non-disaster (low background) | IRR (95% CI) = 1.40 (0.50-4.20) males<br>IRR (95% CI) = 1.10 (0.20-5.30) females |
|  | **Vasiliu et al. 2006** | Males (n=688)<br>Females (n=696) | T2D | Total PCBs | Disaster (contaminated animal feed) | IDR (95% CI) = 1.74 (0.91-3.34) males<br>IDR (95% CI) = 2.33 (1.25-4.34) females |
|  | **Wang et al. 2008** | Males (n=307)<br>Females (n=441) | T2D | Mixed DL-PCBs | Disaster (contaminated rice-oil bran) | OR (95% CI) = 1.7 (0.7-4.3) males<br>OR (95% CI) = 4.6 (1.9-11.4) females |
| **Male-only studies (7 articles)** | **Cappelletti et al. 2016** | n=363 | T2D | PCDD/Fs | Non-disaster (high background) | RR (95% CI) = 2.39 (1.67-3.41) |
|  | **Kang et al. 2006** | n=2,927 | T2D | TCDD | Non-disaster (high background) | OR (95% CI) = 1.49 (1.10-2.02) |
|  | **Kim et al. 2003** | n=1,378 | T2D | TCDD | Non-disaster (high background) | OR (95% CI) = 2.69 (1.09-6.67) |
|  | **Michalek and Pavuk 2008** | n=2,469 | T2D | TCDD | Non-disaster (high background) | RR (95% CI) = 1.58 (1.12-2.24) |
|  | **Persky et al. 2012** | n=63 | T2D | DL-PCBs | Non-disaster (high background) | OR (95% CI) = 2.7 (1.3-5.8) |
|  | **Steenland et al. 2001** | n=2,759 | T2D | TCDD | Non-disaster (high background) | OR (95% CI) = 3.21 (1.81-5.72) |
|  | **Yamamoto et al. 2015** | n=678 | T2D | PCDD/Fs | Non-disaster (high background) | OR (95% CI) = 4.98 (1.17-21.17) |
| **Female-only studies (3 articles)** | **Berg et al. 2021** | n=88 | T2D | DL-PCBs | Non-disaster (low background) | OR (95% CI) = 2.24 (1.0-5.0) |
|  | **Rylander et al. 2015** | n=212 | T2D | PCB-118 | Non-disaster (low background) | OR (95% CI) = 1.55 (0.41-5.86) |
|  | **Zong et al. 2018** | n= 1,586 | T2D | DL-PCBs | Non-disaster (low background) | OR (95% CI) = 1.36 (1.01-1.84) |

### 2.3 Meta-analysis strategy

All forest plots included in the meta-analysis were accompanied with *I*^*2*^-statistics to explore for heterogeneity (Borenstein et al. 2009). All analyses were performed using a random-effects model with OR/RR/IDR summary measures and 95% confidence intervals (CIs). We first performed grouped analysis combining both sexes in all stratified and non-stratified studies. Then, sex-specific associations between T2D incidence and contaminant exposure were evaluated using sub-group analysis via forest plot for male and female populations separately. Lastly, to accurately assess sex-specific susceptibility to T2D within a population, we modeled the difference in the measures of association [Female (OR)] – [Male (OR)] in each sex-stratified study to generate a summary risk difference (RD). Standard errors were calculated based on the 95% CIs from each study.

We also further investigated heterogeneity in the measures of association across mode of exposure (non-disaster versus disaster), type of pollutant, and geographical location (continent). “Non-disaster exposure” was defined to include long-term background low-level exposure typical of the general population (for example through consumption of high fat animal products), chronic “moderate-level” residential exposure in a contaminated region, or long-term high-level occupational exposure (typically military or industrial). “Disaster-exposure” was defined as abrupt, high-level exposure from sudden release of pollutant(s), as through industrial accident or food contamination.

We examined the contribution of publication bias and small study size within the available literature with funnel plots and Egger’s tests (Borenstein et al. 2009) (**Supplementary Figures 1-3**). All analyses were performed using the STATA(SE) 16 statistical software (StataCorp. 2019. *Stata Statistical Software: Release 16*. College Station, TX: StataCorp LLC)

## 3. Results

### 3.1 Summary of articles

Our literature search identified 863 articles, of which 81 met our inclusion criteria (**Figure 2**). Studies were excluded based on the following criteria: (1) studies that did not examine POPs (528 articles); (2) studies that did not examine dioxins or DL POPs (64 articles); (3) studies that did not examine T2D or related metabolic outcomes (105 articles); (4) animal studies or early life exposure studies (37 articles); (5) duplicate studies (48 articles). The age range of study populations in the 81 included studies was >20-59 years (**Supplementary Table 2**).

**Figure 2.**
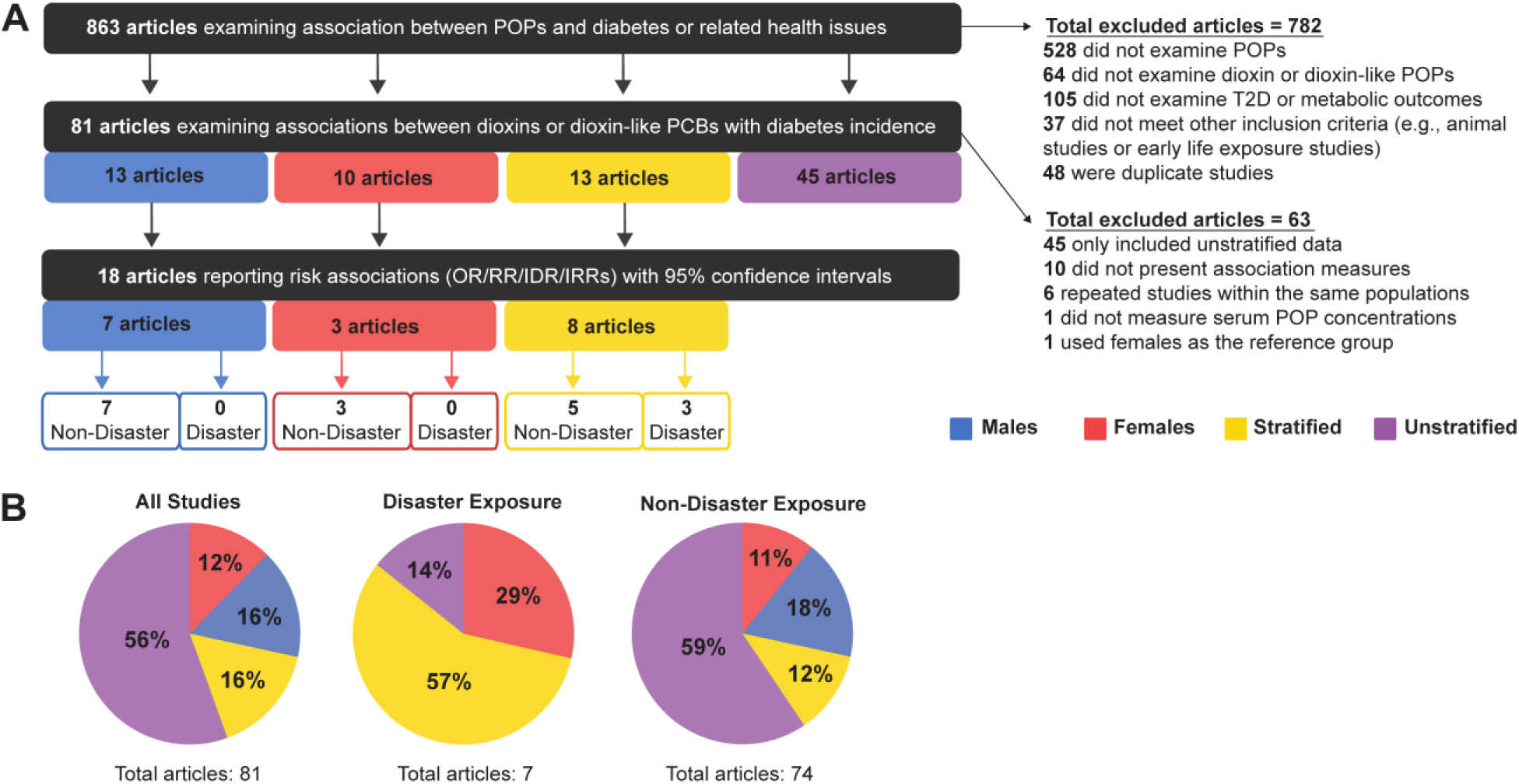
(**A)** Summary of inclusion and exclusion criteria for articles analysed in this meta-analysis, and (**B)** the proportion of studies that were sex-stratified, sex-unstratified, or considered male-only or female-only populations.

Of the 81 articles that initially met our inclusion criteria, the majority (56%, 45 articles) examined sex-unstratified data (i.e. combined male and female) and only 13 studies stratified data by sex (**Figure 2**). There were also 13 studies with data from males only and 10 studies with data from females only (**Figure 2A**). We proceeded to perform our meta-analysis on the 18 articles that either stratified data by sex or reported data from 1 sex and also reported risk association values (Bertazzi et al. 2001; Han et al. 2020; Huang et al. 2015; 2017; Silverstone et al. 2012; Turyk et al. 2009; Vasiliu et al. 2006; Wang et al. 2008; Cappelletti et al. 2016; Kang et al. 2006; Kim et al. 2003; Michalek and Pavuk 2008; Persky et al. 2012; K. Steenland et al. 2001; Yamamoto et al. 2015; Berg et al. 2021; Rylander et al. 2015; Zong et al. 2018). Studies were excluded from meta-analysis if they (1) presented data that was not stratified by sex (45 articles); (2) did not report measures of association (i.e., OR/RR/IDR) (10 articles); (3) repeated measures within an already studied population (of these, only the most recent study was included) (6 articles); (4) did not determine exposure by serum pollutant concentrations (1 article); and (5) did not use an appropriate reference group (1 article) (**Figure 2A**). All studies reported clinical or self-reported T2D or metabolic syndrome diagnosis; most studies of the latter category medically verified self-reported cases. Publication bias using funnel plots and Egger’s test was found to be significant for all studies (p = 0.049) (**Supplementary Figure 1**). However, this significance was driven by the female only data (p = 0.025) (**Supplementary Figure 2**). Male only data showed no publication bias (**Supplementary Figure 3**).

The final 18 studies were subsequently categorized by mode of chemical exposure. Interestingly, of the studies conducted on disaster-exposed populations, 57% were stratified by sex, 29% contained female-only data, 14% were unstratified and none examined male-only data (**Figure 2B**). In contrast, for studies examining non-disaster exposure to dioxin or DL-PCBs, the majority reported sex-unstratified data (59%), followed by male-only data as the second most predominant (18%), and sex-stratified or female-only data composing only 12% and 11% of studies, respectively (**Figure 2B**). Of the 18 articles included in our meta-analysis, 15 articles were non-disaster exposure (7 male-only, 5 stratified, 3 female-only) and 3 articles were disaster exposure, all of which reported sex-stratified data (**Figure 2**).

### 3.2 Both sexes show a significant association between pollutant exposure and T2D

When examining pooled data from all 18 studies, we found that both sexes showed a significant association between pollutant exposure and T2D (**Figure 3, Table 2**). Males showed a 1.95x increased summary risk between exposed and reference populations (95% CI = 1.56-2.43) and females showed a 1.78x increased risk (95% CI = 1.37-2.31) relative to control populations (**Figure 3**). Interestingly, variance due to heterogeneity was significant between the sexes (I^2^ = 51.9%, p = 0.001), indicating that additional variables contribute to determining this association. There was also no significant difference between the sexes when we examined the calculated RD within studies that reported sex-stratified data (RD = 1.18, 95% CI = 0.75-1.87) (**Figure 4)**.

**Figure 3.**
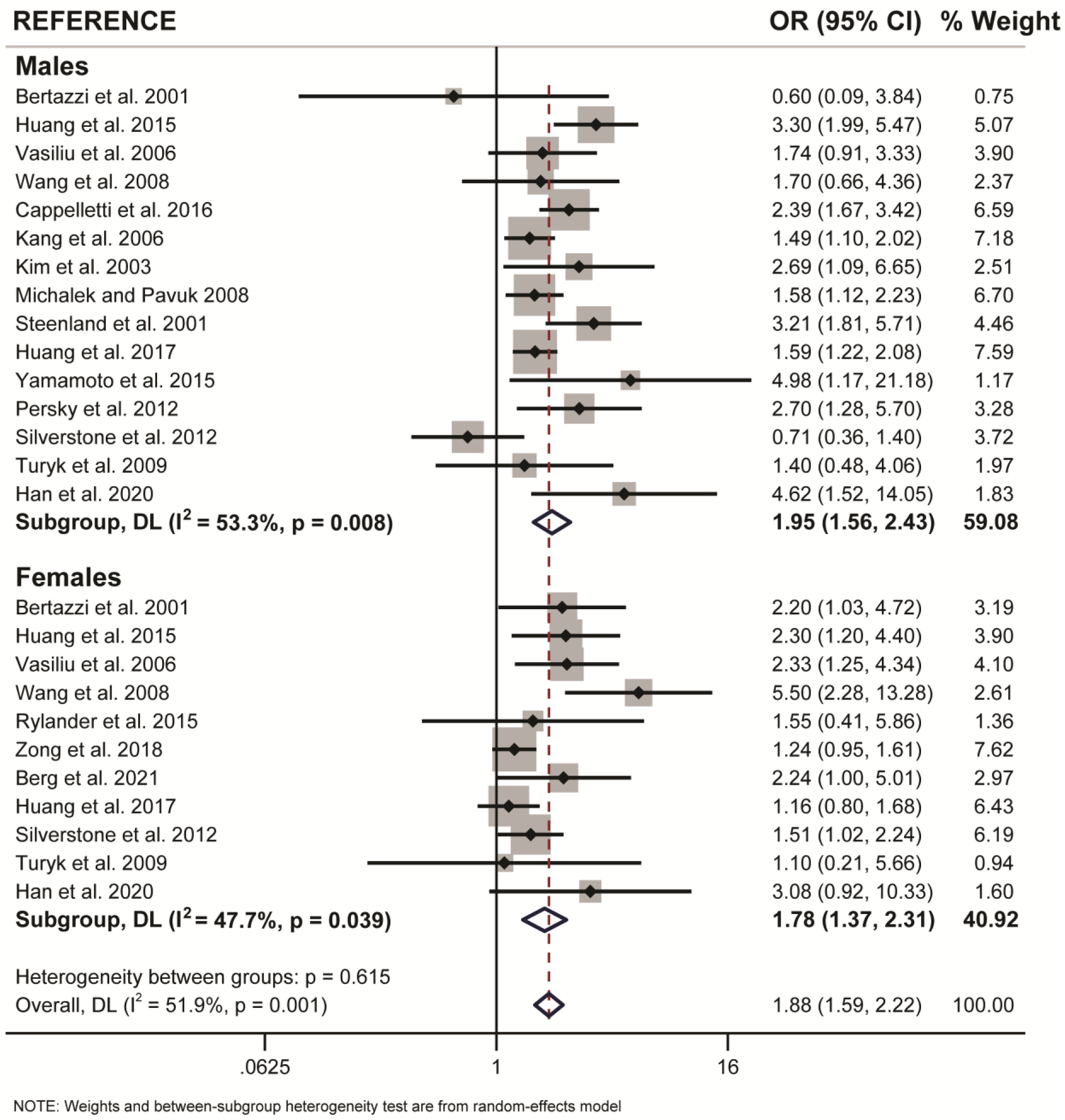
Forest plot of all studies included in meta-analysis, separated by sex. Summary effect measures are significant for both males and females.

**Table 2.**
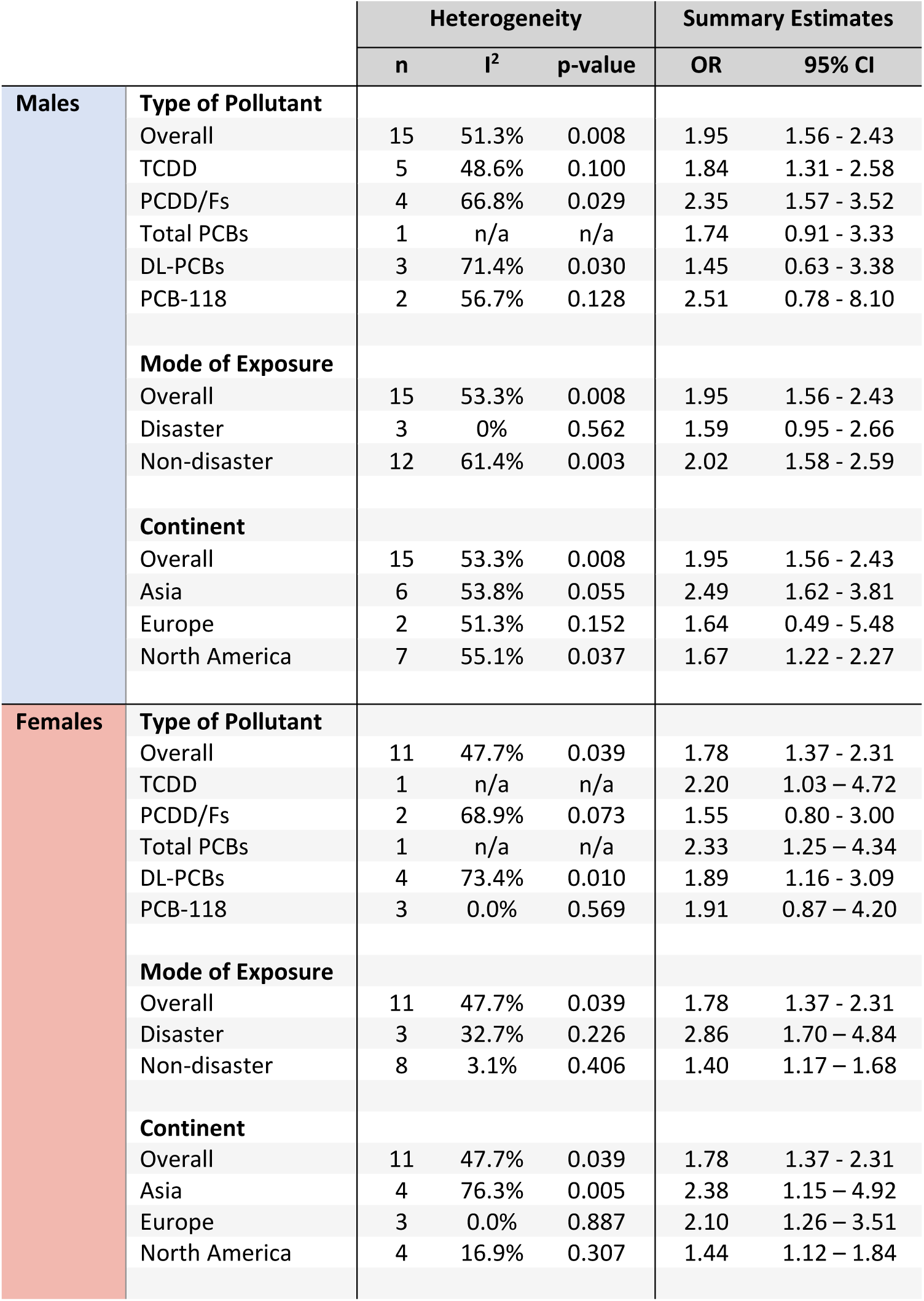
“Male Only” and “Female Only” subgroup analyses of associations between dioxin/DL chemical exposure and T2D outcome. **Note:** “n” = number of entries included in each analysis out of all studies.

|  |  | Heterogeneity |  |  | Summary Estimates |  |
| --- | --- | --- | --- | --- | --- | --- |
|  |  | n | I<sup>2</sup> | p-value | OR | 95% CI |
| Males | **Type of Pollutant** |  |  |  |  |  |
|  | Overall | 15 | 51.3% | 0.008 | 1.95 | 1.56 - 2.43 |
|  | TCDD | 5 | 48.6% | 0.100 | 1.84 | 1.31 - 2.58 |
|  | PCDD/Fs | 4 | 66.8% | 0.029 | 2.35 | 1.57 - 3.52 |
|  | Total PCBs | 1 | n/a | n/a | 1.74 | 0.91 - 3.33 |
|  | DL-PCBs | 3 | 71.4% | 0.030 | 1.45 | 0.63 - 3.38 |
|  | PCB-118 | 2 | 56.7% | 0.128 | 2.51 | 0.78 - 8.10 |
|  | **Mode of Exposure** |  |  |  |  |  |
|  | Overall | 15 | 53.3% | 0.008 | 1.95 | 1.56 - 2.43 |
|  | Disaster | 3 | 0% | 0.562 | 1.59 | 0.95 - 2.66 |
|  | Non-disaster | 12 | 61.4% | 0.003 | 2.02 | 1.58 - 2.59 |
|  | **Continent** |  |  |  |  |  |
|  | Overall | 15 | 53.3% | 0.008 | 1.95 | 1.56 - 2.43 |
|  | Asia | 6 | 53.8% | 0.055 | 2.49 | 1.62 - 3.81 |
|  | Europe | 2 | 51.3% | 0.152 | 1.64 | 0.49 - 5.48 |
|  | North America | 7 | 55.1% | 0.037 | 1.67 | 1.22 - 2.27 |
| Females | **Type of Pollutant** |  |  |  |  |  |
|  | Overall | 11 | 47.7% | 0.039 | 1.78 | 1.37 - 2.31 |
|  | TCDD | 1 | n/a | n/a | 2.20 | 1.03 - 4.72 |
|  | PCDD/Fs | 2 | 68.9% | 0.073 | 1.55 | 0.80 - 3.00 |
|  | Total PCBs | 1 | n/a | n/a | 2.33 | 1.25 - 4.34 |
|  | DL-PCBs | 4 | 73.4% | 0.010 | 1.89 | 1.16 - 3.09 |
|  | PCB-118 | 3 | 0.0% | 0.569 | 1.91 | 0.87 - 4.20 |
|  | **Mode of Exposure** |  |  |  |  |  |
|  | Overall | 11 | 47.7% | 0.039 | 1.78 | 1.37 - 2.31 |
|  | Disaster | 3 | 32.7% | 0.226 | 2.86 | 1.70 - 4.84 |
|  | Non-disaster | 8 | 3.1% | 0.406 | 1.40 | 1.17 - 1.68 |
|  | **Continent** |  |  |  |  |  |
|  | Overall | 11 | 47.7% | 0.039 | 1.78 | 1.37 - 2.31 |
|  | Asia | 4 | 76.3% | 0.005 | 2.38 | 1.15 - 4.92 |
|  | Europe | 3 | 0.0% | 0.887 | 2.10 | 1.26 - 3.51 |
|  | North America | 4 | 16.9% | 0.307 | 1.44 | 1.12 - 1.84 |

**Figure 4.**
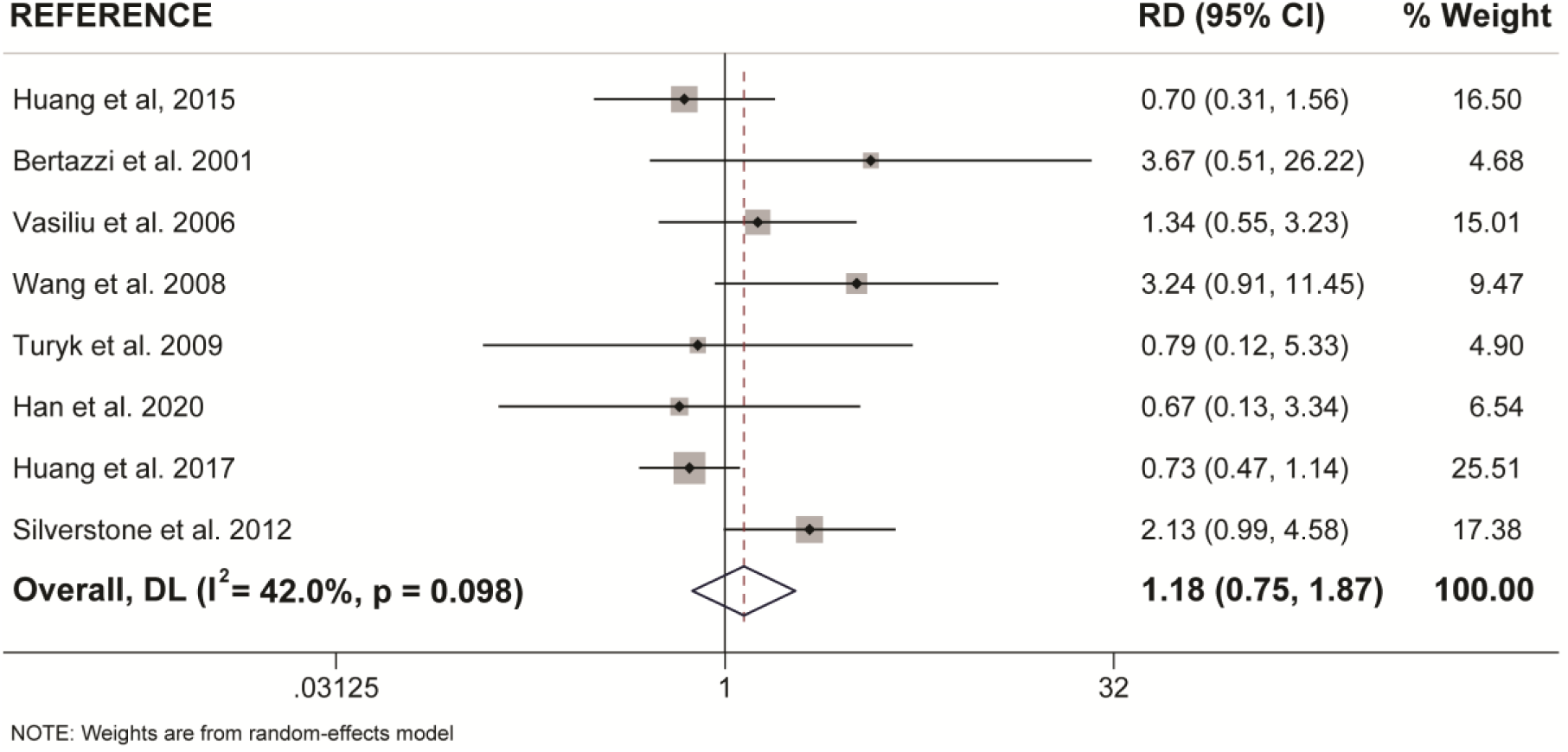
Forest plot of risk difference for pollutant exposure and diabetes incidence between males and females from the same study population (only sex-stratified studies included). Summary measures were generated from the difference in summary effect measures between the sexes (Females - Males).

We next looked for sex differences when studies were further subcategorized by population location or type of pollutant (**Table 2**). There were no clear sex differences within either of these subcategories, with the exception of DL-PCBs, which showed a significant association with T2D in females (OR = 1.89, 95% CI = 1.16-3.09; 4 studies), but not males (OR = 1.45, 95% CI = 0.63-3.38; 3 studies) (**Table 2**). However, most of these subcategories had too few studies to rule out potential sex differences.

### 3.3 Sex differences when considering mode of exposure

Our meta-analysis revealed clear sex-specific associations when we categorized studies by mode of exposure (i.e., disaster versus non-disaster). Males exposed to pollutants via non-disaster showed a significant 2.02x increased risk for T2D incidence relative to control populations (95% CI = 1.58-2.59), compared to a non-significant 1.59x increased risk when exposed via disaster (95% CI = 0.95-2.66) (**Figure 5A, Table 2**). Females show the opposite trend, with a modest 1.40x increased risk associated with non-disaster exposure (95% CI = 1.17-1.68) compared to a pronounced 2.86x increased risk associated with disaster exposure (95% CI = 1.70-4.84) (**Figure 5B, Table 2**). Therefore, in disaster exposure conditions, the increased risk for T2D is primarily driven by a significant increase in the OR for females but not males.

**Figure 5.**
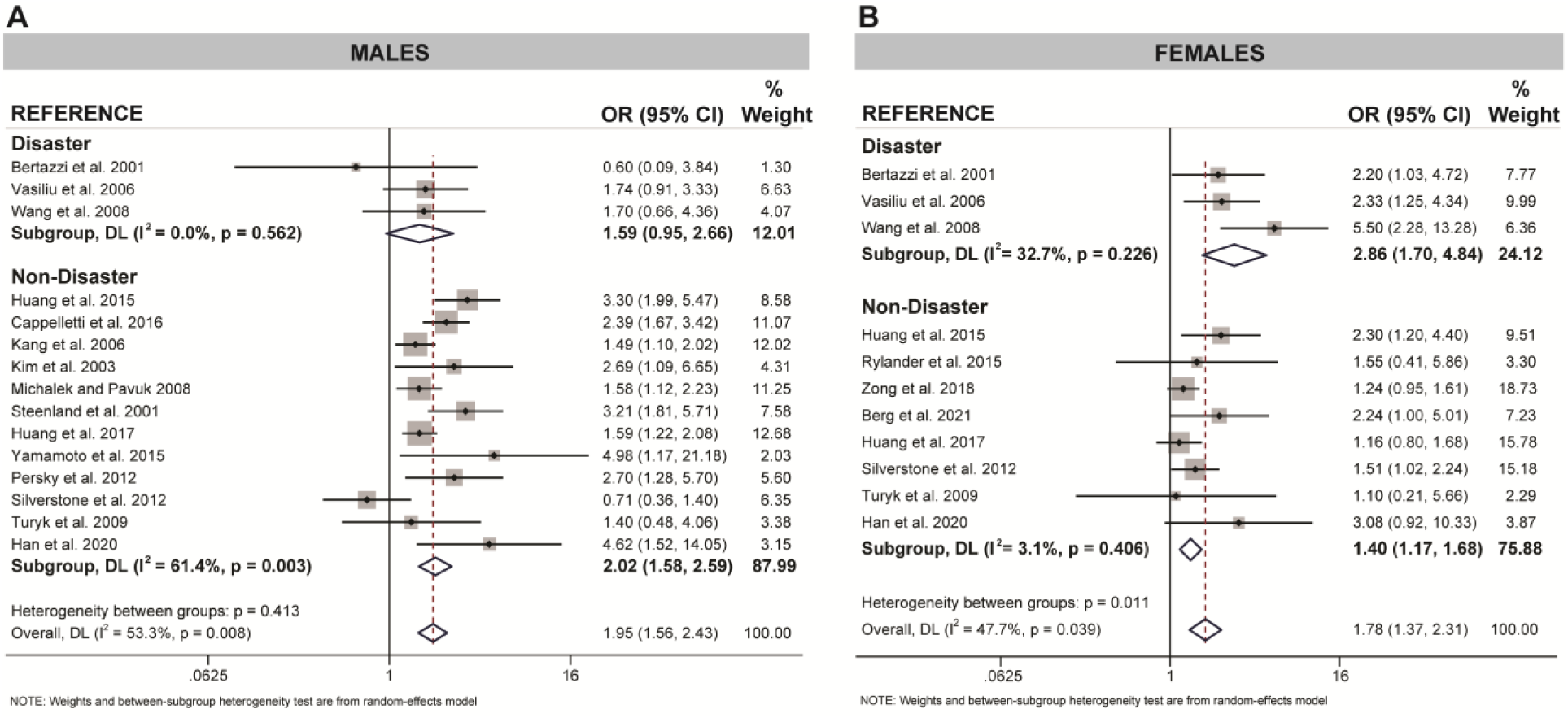
Forest plot of association between pollutant exposure and diabetes incidence with separate analysis for disaster and non-disaster exposure in (**A)** males and **(B)** females.

To further investigate these sex differences, we performed a meta-analysis on only sex-stratified studies using the calculated differences in association measures between the sexes within the same population (**Figure 6**). As expected, there was no difference in the risk difference between the sexes when considering non-disaster exposure studies (OR = 0.93, 95% CI = 0.57-1.52). Interestingly, there was a 1.95x increase in risk difference between the sexes when only disaster-exposure studies were analyzed (95% CI = 0.99-3.84) (**Figure 6**).

**Figure 6.**
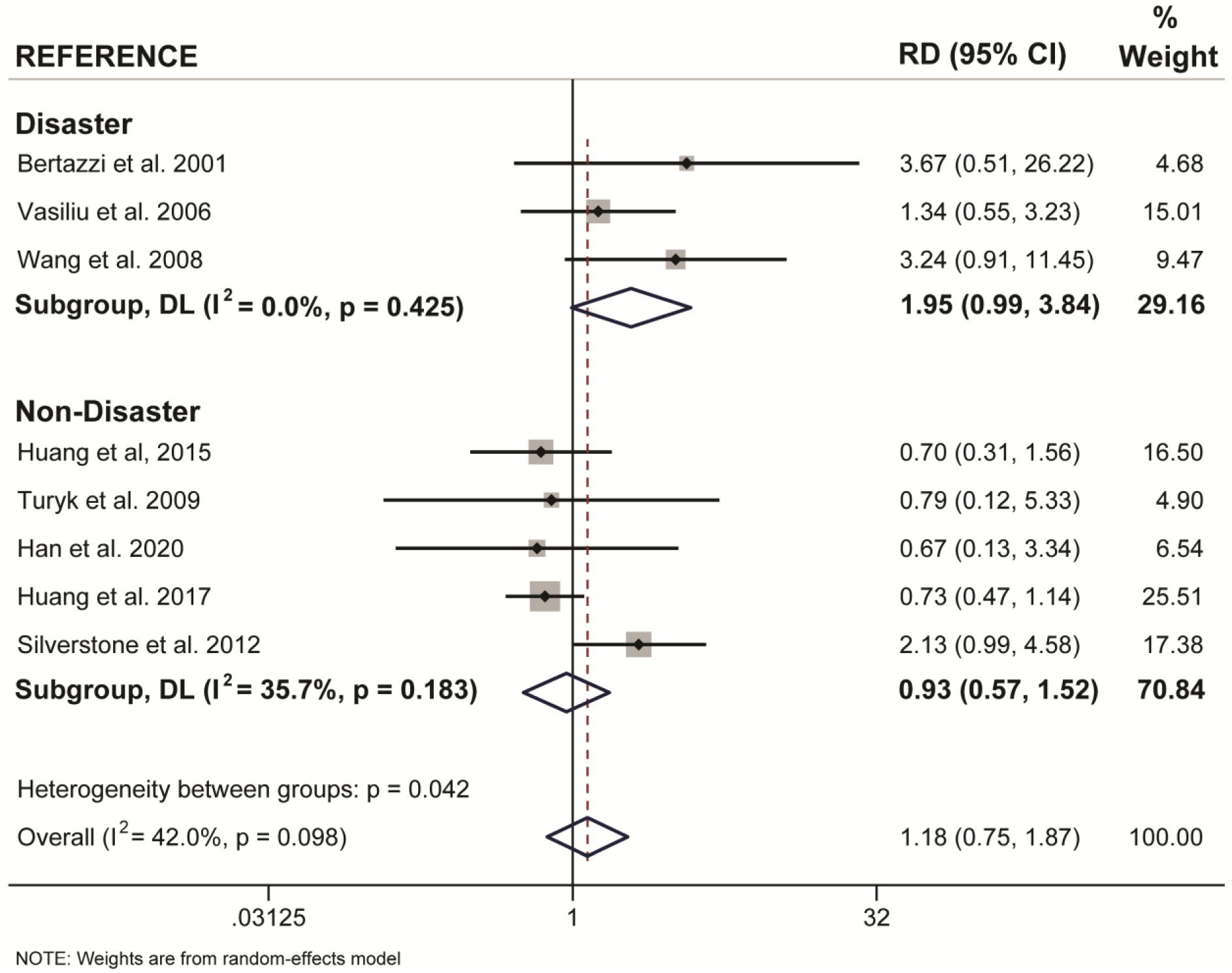
Forest plot of risk difference for disaster versus non-disaster pollutant exposure and diabetes incidence between males and females from the same study population (only sex-stratified studies included). Summary measures were generated from the difference in summary effect measures between the sexes (Females - Males) and stratified by mode of exposure.

## 4. Discussion

A total of 18 articles were included in this meta-analysis to investigate whether there are sex-specific associations between dioxin or DL-PCB exposure and T2D incidence in adult populations. When we considered all studies together, there were no sex differences in this association, but sex differences emerged when data was further stratified by mode of exposure. We found that the strength of association between exposure to dioxin/DL-PCBs via disaster and incident diabetes was stronger in females than males. Importantly, this sex difference was consistent when we calculated the summary OR for females versus males as well as the risk difference within matched populations. To our knowledge, mode of exposure has not previously been considered when examining sex differences in epidemiology literature on pollutant exposure.

Our review of the epidemiological literature revealed important gaps that limited our meta-analysis of sex-specific associations between pollutant exposure and diabetes. Sex-stratified studies accounted for only 16% of the literature examining links between diabetes and dioxin or DL-PCB exposure. Furthermore, most studies (56%) in the general population did not stratify data by sex, which limited our analysis of non-disaster exposure. Additional research in sex-stratified populations is essential to confirm the sex differences revealed by our meta-analysis in disaster-exposed populations and to elucidate potential sex differences in the general population. Female only data showed publication bias, whereas male only data did not, which strengthens our assertion that sex-differences need to be considered in future epidemiological studies in the general population. It would also be of interest to understand if the pollutant burden differs between sexes following disaster-exposure, as this could contribute to the sex differences found in our meta-analyses. A more comprehensive meta-analysis that also considers other POPs, such as organochlorine pesticides or perfluorinated chemicals, could facilitate a more complete picture of the literature and identify potential sex-specific associations. In addition, carefully controlled studies in model systems can provide valuable insight into potential mechanistic links, different modes of pollutant exposure (e.g. chronic low-dose versus single high-dose), and sex-specific diabetes pathogenesis.

Despite limitations in sex-stratified epidemiological literature, we noted parallels between the human and rodent data that strengthen our hypothesis that females may be more prone to dioxin/DL pollutant-induced diabetes than males. For example, in human populations with disaster-exposure to dioxins/DL-PCBs, females had a higher risk for T2D than males. The rodent study that best models a disaster exposure scenario is a single, high-dose TCDD exposure protocol, in which TCDD-exposed female but not male mice developed transient hyperglycemia compared to vehicle-exposed controls (Hoyeck et al. 2020). To mimic the background-level exposure that humans experience in non-disaster settings, a repeated low-dose chemical exposure protocol in rodents is often used. Our lab reported that low-dose TCDD exposure for 12 weeks did not disrupt glucose homeostasis in male or female mice fed a chow diet (Matteo et al. 2020). However, TCDD accelerated the onset of high fat diet-induced hyperglycemia in female mice but not male mice (Matteo et al. 2020). The epidemiology data in non-disaster cohorts showed a consistent increased risk of diabetes incidence in both males and females but these studies do not consider BMI or diet composition, which may be important covariates contributing to sex-specific associations. Another consideration is that low-dose TCDD exposure for 12 weeks in mice is far from the life-long exposure experienced by humans. So while male mice did not develop hyperglycemia in this timeframe, longer-term studies are certainly warranted given the clear epidemiological association between background level TCDD exposure and increased diabetes incidence in males.

One potential mechanism for the effects of dioxin/DL-PCB exposure on glucose homeostasis is AhR activation in metabolic tissues, including pancreatic islets (Ibrahim et al. 2020). Interestingly, AhR upregulates CYP enzyme isoforms in a sex-specific manner (Kwekel et al. 2010; L. Yang and Li 2012) and several studies have reported higher levels of CYP enzymes, including CYP1A1, CYP1A2, CYP1B1 in females compared to males in mice, pigs, and humans (Skaanild and Friis 1999; Finnström et al. 2002; Lu et al. 2013). In addition, Roh et al. (2015) reported that in humans, females had significantly higher AhR ligand activity (including both exogenous and endogenous ligands) than males. There was also a significant association between increased serum AhR ligand activity and T2D (Roh et al. 2015). The AhR pathway is involved in multiple essential cellular functions, including xenobiotic metabolism, cell cycle, inflammation, circadian rhythm, adhesion and migration, cellular plasticity, and estrogen receptor signaling (Larigot et al. 2018; Nebert 2017; Quintana and Sherr 2013; Bock 2018; Kung, Murphy, and White 2009; Anderson et al. 2013; Lu et al. 2013; Swedenborg and Pongratz 2010). Exploring potential mechanisms underlying sex differences in AhR activation and CYP enzyme expression are beyond the scope of this article, but deserve further study.

Age of exposure is an important covariate that we could not control for in our meta-analysis, but we limited our inclusion criteria to studies on adult populations to minimize variability. Childhood exposure to dioxins and DL-PCBs is associated with myriad complications that can present at birth (Tawara et al. 2009; Nghiem et al. 2019) and childhood (Nguyen et al. 2018; Nishijo et al. 2012; Tai et al. 2016; Tran et al. 2016; Pham et al. 2020; Z. Wang et al. 2019; Ames et al. 2019), and subsequently impact adult health. There are also interesting sex-specific effects reported in these populations (Nishijo et al. 2012; Nguyen et al. 2018; Tai et al. 2016; Pham et al. 2020; Z. Wang et al. 2019; Ames et al. 2019), further emphasizing the need for sex-stratified data in the epidemiology literature. However, the current study was intentionally designed to focus on the association between adult exposure and adult presentation of T2D.

In summary, this review compared articles that examined associations between T2D incidence and pollutant exposure from either a disaster or non-disaster setting, which included high occupational background exposure and low ubiquitous background exposure. Females showed statistically significant associations between dioxin/DL-PCB exposure and increased diabetes risk under disaster conditions, whereas males did not. More epidemiological studies with sex-stratified data are needed to confirm this observation and further investigate potential sex differences within the general population. Collectively, this work will help to inform legislation and policy-makers on taking measures towards pollutant control.

## Data Availability

All data generated from the meta-analysis and literature review are provided in the manuscript and supplementary document. Additional information can be provided upon request.

## Acknowledgements

We thank Myriam Hoyeck and Erin Mulvihill for thoughtful discussions of the literature and assistance with literature searches and article classification. We thank Kayleigh Rick for her assistance with the literature search. We thank Geronimo Matteo for his assistance in article preparation. This research was supported by a Canadian Institutes of Health Research (CIHR) Project Grant (#PJT-2018-159590).

