## Supplementary Tables and Figures for "Sex-specific associations between type 2 diabetes incidence and exposure to dioxin and dioxin-like pollutants: a meta-analysis"

Supplementary Figures

**Supplementary Table 1.** Literature search terms

| | |
|---|---|
| Database: Pubmed (May 17, 2021) <2020 | |
| Search Strategy: | |
| "Review" [Publication Type], Filter: Humans | |
| 1 | "Diabetes Mellitus, Type 2"[Mesh] OR "Diabetes Mellitus"[Mesh:NoExp] OR "Hyperglycemia"[Mesh] OR "Prediabetic State"[Mesh] OR "Metabolic Syndrome"[Mesh] OR diabetes[Title/Abstract] OR hyperglycemia[Title/Abstract] OR pre-diabet*[Title/Abstract] OR prediabet*[Title/Abstract] OR dysglycemia[Title/Abstract] OR "glucose intolerance"[Title/Abstract] OR "metabolic syndrome"[Title/Abstract] OR "insulin resistance syndrome"[Title/Abstract] OR "dysmetabolic syndrome"[Title/Abstract] OR "reaven syndrome x"[Title/Abstract] OR "cardiometabolic syndrome"[Title/Abstract] |
| 2 | "Benzofurans"[Mesh] OR benzofuran*[Title/Abstract] OR BFPA[Title/Abstract] OR PCDF[Title/Abstract] OR PCDFs[Title/Abstract] OR Amiodarone[Title/Abstract] OR Dronedarone[Title/Abstract] OR Benzbromaron*[Title/Abstract] OR Cantharid*[Title/Abstract] OR Citalopram[Title/Abstract] OR Fluorescamine[Title/Abstract] OR Fura-2[Title/Abstract] OR Griseofulvin[Title/Abstract] OR Pterocarpans[Title/Abstract] OR Vilazodone Hydrochloride[Title/Abstract] OR Amiohexal[Title/Abstract] OR Aratac[Title/Abstract] OR Tachydaron[Title/Abstract] OR Corbionax[Title/Abstract] OR Cordarone[Title/Abstract] OR Amiodarex[Title/Abstract] OR Trangorex[Title/Abstract] OR Kordaron[Title/Abstract] OR Cordarex[Title/Abstract] OR L-3428[Title/Abstract] OR "L 3428"[Title/Abstract] OR L3428[Title/Abstract] OR Ortacrone[Title/Abstract] OR Rytmarone[Title/Abstract] OR "SKF 33134-A"[Title/Abstract] OR "SKF 33134 A"[Title/Abstract] OR "SKF 33134A"[Title/Abstract] OR Amiobeta[Title/Abstract] OR Braxan[Title/Abstract] OR Multaq[Title/Abstract] OR "SR 33589B"[Title/Abstract] OR "SR 33589"[Title/Abstract] OR Narcaricin[Title/Abstract] OR Desuric[Title/Abstract] OR Urinorm[Title/Abstract] OR Acifugan[Title/Abstract] OR Besuric[Title/Abstract] OR Cytalopram[Title/Abstract] OR Seropram[Title/Abstract] OR Escitalopram[Title/Abstract] OR Celexa[Title/Abstract] OR Lu-10-171[Title/Abstract] OR Lu10171[Title/Abstract] OR "Escitalopram Oxalate"[Title/Abstract] OR Lexapro[Title/Abstract] OR Fluram[Title/Abstract] OR Grisefuline[Title/Abstract] OR Gris-PEG[Title/Abstract] OR "Gris PEG"[Title/Abstract] OR GrisPEG[Title/Abstract] OR Grisactin[Title/Abstract] OR Fulvicin-U-F[Title/Abstract] OR "Fulvicin U F"[Title/Abstract] OR FulvicinUF[Title/Abstract] OR "Grifulvin V"[Title/Abstract] OR Viibryd[Title/Abstract] OR "EMD 68843"[Title/Abstract] OR EMB-68843[Title/Abstract] OR "EMB 68843"[Title/Abstract] OR EMB68843[Title/Abstract] OR Vilazodone[Title/Abstract] OR fluspidine[Title/Abstract] OR mecinarone[Title/Abstract] OR diMeMEBFE[Title/Abstract] OR suillusin[Title/Abstract] OR rocaglaol[Title/Abstract] OR "1,3-dihydroisobenzofuran"[Title/Abstract] OR phthalane[Title/Abstract] OR 1-ACSFC[Title/Abstract] OR "5-(fluoromethoxy)-MOPSBCC"[Title/Abstract] OR |

| | |
|---|---|
| | ABT239[Title/Abstract] OR ABT-239[Title/Abstract] OR "MDHMDBP cpd"[Title/Abstract] OR naltriben[Title/Abstract] OR SBFI[Title/Abstract] OR benzofury[Title/Abstract] OR gymnastone[Title/Abstract] OR KSCM[Title/Abstract] OR TAK-937[Title/Abstract] OR SNU-0039[Title/Abstract] OR 2-methyl-MPBO[Title/Abstract] OR HS-113[Title/Abstract] OR fasiglifam[Title/Abstract] OR artonitidin[Title/Abstract] OR TC5619[Title/Abstract] OR TC-5619[Title/Abstract] OR spirolaxine[Title/Abstract] OR 5-acetyl-DHMBABE[Title/Abstract] OR "5,6-DMEBF"[Title/Abstract] OR Thunbergol[Title/Abstract] OR Hedysarumine[Title/Abstract] OR 2-ethyl-CTDBCA[Title/Abstract] OR N-PBuBzFCMA[Title/Abstract] OR "4-(FPHMB)-BFPCP"[Title/Abstract] OR SB331750[Title/Abstract] OR SB-331750[Title/Abstract] OR KB130015[Title/Abstract] OR KB-130015[Title/Abstract] OR 2-DEAPPB[Title/Abstract] OR 3-TDEAPB[Title/Abstract] OR MEN-1162*[Title/Abstract] OR ethofumesate[Title/Abstract] OR motegrity[Title/Abstract] OR resotran[Title/Abstract] OR resotrans[Title/Abstract] OR R 093877[Title/Abstract] OR R093877[Title/Abstract] OR Resolor[Title/Abstract] OR LY320135[Title/Abstract] OR LY-320135[Title/Abstract] OR MDL-74180[Title/Abstract] OR U-63557A[Title/Abstract] OR afabicin[Title/Abstract] OR N3RQ532IUT[EC/RN Number] OR JQZ1L091Y2[EC/RN Number] OR 4POG0RL69O[EC/RN Number] OR IGL471WQ8P[EC/RN Number] OR 0DHU5B8D6V[EC/RN Number] OR 38183-12-9[EC/RN Number] OR TSN3DL106G[EC/RN Number] OR 32HRV3E3D5[EC/RN Number] OR U8HTX2GK8J[EC/RN Number] OR 150563-61-4[EC/RN Number] OR LK6946W774[EC/RN Number] OR 140129-67-5[EC/RN Number] OR 138663-21-5[EC/RN Number] OR 86H6B395PI[EC/RN Number] OR 111555-58-9[EC/RN Number] OR 124549-08-2[EC/RN Number] OR 285VE60914[EC/RN Number] OR 3051U1Z8KV[EC/RN Number] OR 0A09IUW5TP[EC/RN Number] OR CJY03984CT[EC/RN Number] OR 133806-61-8[EC/RN Number] OR 133806-60-7[EC/RN Number] OR 124487-65-6[EC/RN Number] OR 94F21T5G3C[EC/RN Number] OR 94663-91-9[EC/RN Number] OR KX4D9BZA6X[EC/RN Number] OR 53737-95-4[EC/RN Number] OR 5196-74-7[EC/RN Number] OR 1EVW48QLUN[EC/RN Number] OR 77828-26-3[EC/RN Number] OR DMM8663H2R[EC/RN Number] OR 2R6NTQ7Y6G[EC/RN Number] OR N917K7H53K[EC/RN Number] OR 140853-59-4[EC/RN Number] OR 140853-58-3[EC/RN Number] OR 7B1550J80K[EC/RN Number] |
| 3 | "Dioxins and Dioxin-like Compounds"[Mesh:NoExp] OR "Dioxins"[Mesh] OR "Polychlorinated Biphenyls"[Mesh] OR "persistent organic pollutants"[Title/Abstract] OR POPs[Title/Abstract] OR dibenzo-p-dioxin*[Title/Abstract] OR dibenzodioxin*[Title/Abstract] OR "polychlorinated biphenyl"[Title/Abstract] OR "polychlorinated biphenyls"[Title/Abstract] OR PCDD[Title/Abstract] OR PCDDs[Title/Abstract] OR PBDD[Title/Abstract] OR PBDDs[Title/Abstract] OR PXDD[Title/Abstract] OR PXDDs[Title/Abstract] OR Dioxin*[Title/Abstract] OR Polychlorodibenzodioxin*[Title/Abstract] OR dichlorodibenzodioxin*[Title/Abstract] OR trichlorodibenzodioxin*[Title/Abstract] OR tetrachlorodibenzodioxin*[Title/Abstract] OR pentachlorodibenzodioxin*[Title/Abstract] OR hexachlorodibenzodioxin*[Title/Abstract] OR heptachlorodibenzodioxin*[Title/Abstract] OR octochlorodibenzodioxin*[Title/Abstract] OR Polychlorodibenzo-p-dioxin* [Title/Abstract] OR dichlorodibenzo-p-dioxin*[Title/Abstract] OR trichlorodibenzo-p- |

| | |
|---|---|
| | dioxin*[Title/Abstract] OR tetrachlorodibenzo-p-dioxin*[Title/Abstract] OR pentachlorodibenzo-p-dioxin*[Title/Abstract] OR hexachlorodibenzo-p-dioxin*[Title/Abstract] OR heptachlorodibenzo-p-dioxin*[Title/Abstract] OR octochlorodibenzo-p-dioxin*[Title/Abstract] OR "Agent orange"[Title/Abstract] OR TCDD[Title/Abstract] OR HxCDD[Title/Abstract] OR HpCDD[Title/Abstract] OR OCDD[Title/Abstract] OR TCDDs[Title/Abstract] OR HxCDDs[Title/Abstract] OR HpCDDs[Title/Abstract] OR OCDDs[Title/Abstract] OR Polychlorobiphenyl*[Title/Abstract] OR PCB[Title/Abstract] OR PCBs[Title/Abstract] OR Aroclor*[Title/Abstract] OR Chlorodiphenyl*[Title/Abstract] OR dichlorobiphenyl*[Title/Abstract] OR trichlorobiphenyl*[Title/Abstract] OR tetrachlorobiphenyl*[Title/Abstract] OR pentachlorobiphenyl*[Title/Abstract] OR hexachlorobiphenyl*[Title/Abstract] OR heptachlorobiphenyl*[Title/Abstract] OR CC0651[Title/Abstract] OR C-0651[Title/Abstract] OR DMPCAP[Title/Abstract] OR DCB[Title/Abstract] OR TCB[Title/Abstract] OR TBP[Title/Abstract] OR HTCBP[Title/Abstract] OR HTClBP[Title/Abstract] OR Delor 103[Title/Abstract] OR Delor-103[Title/Abstract] OR BMSTBP[Title/Abstract] OR BESTBP[Title/Abstract] OR HPCB[Title/Abstract] OR B94L7A4G3D[EC/RN Number] OR HF5S8P28CC[EC/RN Number] OR 5R0UY33JFS[EC/RN Number] OR B94L7A4G3D[EC/RN Number] OR MM6333103R[EC/RN Number] OR 6W0BQ34FSH[EC/RN Number] OR 39227-58-2[EC/RN Number] OR 34465-46-8[EC/RN Number] OR YW59P10266[EC/RN Number] OR TTP086070W[EC/RN Number] OR 25569-80-6[EC/RN Number] OR S0B721XXDK[EC/RN Number] OR 0S2FWS766G[EC/RN Number] OR 62U12S7CJZ[EC/RN Number] OR 35R7J07E3D[EC/RN Number] OR IXL47709VM[EC/RN Number] OR DOF6S5DT2R[EC/RN Number] OR 57308-11-9[EC/RN Number] OR 1433W7U14D[EC/RN Number] OR 37680-65-2[EC/RN Number] OR 1ZNA5F571U[EC/RN Number] OR 3370393855[EC/RN Number] OR 03KW6RT30C[EC/RN Number] OR 35693-92-6[EC/RN Number] OR 98H9867N6L[EC/RN Number] OR 844ODP31Q0[EC/RN Number] OR 38444-84-7[EC/RN Number] OR 66640-67-3[EC/RN Number] OR Y2I6546TMI[EC/RN Number] OR 88966-73-8[EC/RN Number] OR 4RPD28X1JO[EC/RN Number] OR 38444-93-8[EC/RN Number] OR 4470AQR07D[EC/RN Number] OR M8CJF43Q12[EC/RN Number] OR 1EH557950R[EC/RN Number] OR W0250484DW[EC/RN Number] OR 392EXD3SC9[EC/RN Number] OR PD736T0W8F[EC/RN Number] OR LH750S39FX[EC/RN Number] OR 1150VTX8QQ[EC/RN Number] OR F7XE9G6462[EC/RN Number] OR 36559-22-5[EC/RN Number] OR K8L6S986MK[EC/RN Number] OR 104104-34-9[EC/RN Number] OR K2Y92IEV5R[EC/RN Number] OR 60640-55-3[EC/RN Number] OR 94659-41-3[EC/RN Number] OR 66640-68-4[EC/RN Number] OR 68099-35-4[EC/RN Number] OR BT5E4PD553[EC/RN Number] OR TSH69IA9XF[EC/RN Number] OR Z4YYF101N3[EC/RN Number] OR 86S23H0DSX[EC/RN Number] OR 803YVI5BNP[EC/RN Number] OR 3PY435PFIO[EC/RN Number] OR 2B2AQE8U50[EC/RN Number] OR 0S596V3MLH[EC/RN Number] OR 130689-92-8[EC/RN Number] OR 67651-36-9[EC/RN Number] OR 1D11835905[EC/RN Number] OR 0YO8J06WCR[EC/RN Number] OR 97RWU5J04P[EC/RN Number] OR VIF9S75RZV[EC/RN Number] OR VIU6O9X06D[EC/RN Number] OR HD4MZI40CH[EC/RN Number] OR 7Y6JIG1867[EC/RN Number] OR |

| | |
|---|---|
| | P7EUD9W45V[EC/RN Number] OR R9YK3084VS[EC/RN Number] OR T2P1WH546D[EC/RN Number] OR PJ11O9J71Y[EC/RN Number] OR ZRU0C9E32O[EC/RN Number] OR MOA04J1VTW[EC/RN Number] OR 5JNS37UVGI[EC/RN Number] OR 38380-05-1[EC/RN Number] OR 38380-08-4[EC/RN Number] OR 52663-63-5[EC/RN Number] OR 56030-56-9[EC/RN Number] OR 39635-31-9[EC/RN Number] OR 2I988FUS7H[EC/RN Number] OR 52I0CG8IQX[EC/RN Number] OR CIP7BRA48E[EC/RN Number] OR 33091-17-7[EC/RN Number] |

**Supplementary Table 2.** In-depth summary of 18 articles investigating the association between dioxins and/or DL-PCBs and diabetes incidence included in the meta-analysis.

| | Study Years and Authors | Cohort | Study Participants | Median participant age (yr) | Type of Study | Outcome | Type of Exposure | Mean Pollutant Exposure | Adjustments | Results (OR, RR, IDR, IRR) |
|---|---|---|---|---|---|---|---|---|---|---|
| Sex-stratified studies | Bertazzi et al. 2001 | Seveso, Italy cohort (20-year follow-up) | Males (n=22,607) Females (n=22,762) | 50 | Cohort | T2D | Disaster (Plant Explosion) | TCDD<br>Zone A = 443 pg/g<br>Zone B = 87 pg/g<br>Zone R = 15 pg/g | Age, calendar period of data collection, and sex | RR (95% CI) = 0.6(0.1-4.1) males. RR (95% CI) = 2.2(1.0-4.6) females |
| | Han et al. 2020 | Shandong, East China cohort | Males (n=151) Females (n=165) | 50 | Case-control | T2D | Non-disaster (low background) | PCB-118 (0.2 ng/mL) | Age, BMI, total cholesterol and triglycerides | OR (95% CI) = 4.62(1.52-14.06) males.<br>OR (95% CI) = 3.08 (0.92-10.35) females |
| | Huang et al. 2015 | Tainan City, Taiwan cohort | n = 2,898 (total participants) | 52 | Cohort | T2D | Non-disaster (high background) | PCDD/Fs (20-62 pg/g lipid) | Age, BMI, duration or residency in locale | OR (95% CI) = 3.3(2.0-5.5) males. OR (95% CI) = 2.3(1.2-4.4) females |
| | Huang et al. 2017 | Tainan City, Taiwan cohort | Males (n=1,453) Females (n=1,305) | 50 | Cohort | Metabolic Syndrome | Non-disaster (higher background) | PCDD/Fs (770 pg/g lipid males; 675 pg/g lipid females) | Age | OR (95% CI) = 1.59 (1.22-2.08) males. OR (95% CI) = 1.16 (0.8-1.68) females |

| | | | | | | | | | | |
|---|---|---|---|---|---|---|---|---|---|---|
| | Silverstone et al. 2012 | Anniston Health Community Survey | Males (n=153) Female (n=427) | 55 | Cohort | T2D | Non-disaster (high background) | DL-PCBs (9.33 ppb) | Age, BMI, total lipids, race/ethnicity, family history of diabetes, and current medications | OR (95% CI) = 0.71(0.36-1.39) males. OR (95% CI) = 1.51(1.02-2.24) females |
| | Turyk et al. 2009 | Great Lakes Sport Fish Eaters cohort | Males (n=279) Females (n=192) | 52 (males) 47 (females) | Cohort | T2D | Non-disaster (low background) | PCB-118 (2.4 ng/g lipid males; 0.7 ng/g lipid females) | Age and BMI | IRR (95% CI) = 1.40(0.50-4.20) males. IRR (95% CI) = 1.10(0.20-5.30) females |
| | Vasiliu et al. 2006 | Michigan, USA cohort (25-year follow-up) | Males (n=688) Females (n=696) | 45 | Cohort | T2D | Disaster (contaminated animal feed) | Total PCBs (7.0 ppb) | Age, BMI, alcohol consumption, cigarette smoking | IDR (95% CI) =1.74(0.91-3.34) males. IDR (95% CI) = 2.33(1.25-4.34) females |
| | Wang et al. 2008 | Yucheng, Taiwan cohort (24-year follow-up) | Males (n=307) Females (n=441) | 59 (males) 52 (females) | Cohort | T2D | Disaster (contaminated rice-oil bran) | Mixed DL-PCBs (73.3 ppb males; 87.4 ppb females) | Age | OR (95% CI) = 1.7(0.7-4.2) males. OR (95% CI) = 4.6(1.9-11.4) females |
| Male – only studies | Cappelletti et al. 2016 | Italian Electric Arc Furnace (EAF) Workers cohort | n=363 | >20 | Cohort | T2D | Non-disaster (high background) | PCDD/Fs (0.092-0.693 mg/kg dust) | Age | RR (95% CI) = 2.39(1.67-3.41) |
| | Kang et al. 2006 | US Vietnam veterans (Army Chemical Corps) | n=2,927 | 52 | Cohort | T2D | Non-disaster (high background) | TCDD (4.3 ppt) | Age, race/ethnicity, BMI, and current smoking status | OR (95% CI) = 1.49 (1.10-2.02) |

| | | | | | | | | | | |
|---|---|---|---|---|---|---|---|---|---|---|
| | Kim et al. 2003 | Korean Vietnam veteran cohort) | n=1,378 | 55 | Cohort | T2D | Non-disaster (high background) | TCDD (0.62 pg/g) | Age, smoking, alcohol, BMI, education, and marital status | OR (95% CI) = 2.69(1.09-6.67) |
| | Michalek and Pavuk 2008 | US Vietnam Veterans (Operation Ranch Hand) Cohort | n=2,469 | 45 | Case-Control | T2D | Non-disaster (high background) | TCDD (10 ppt) | Age, race/ethnicity, and military occupation | RR (95% CI) 1.58 (1.12-2.24) |
| | Persky et al. 2012 | Utilities Company (EUC) Past Employees (US) Cohort | n=63 | 58 | Cross-sectiona l | T2D | Non-disaster (high background) | DL-PCBs (2.5 ng/g lipid) | Age, BMI and total lipids | OR (95% CI) = 2.7 (1.3-5.8) |
| | Steenland et al. 2001 | US Vietnam Veterans (operation ranch hand) and National Institute for Occupational Safety and Health (NIOSH) workers cohorts) | n=2,759 | 53 | Cohort | T2D | Non-Disaster (High background) | TCDD (69 ppt) | Family history of diabetes, BMI, age, race/ethnicity, current medications, and education | OR (95% CI) = 3.21 (1.81-5.72) |
| | Yamamoto et al. 2015 | Japanese Incinerator Workers cohort | n=678 | 55 | Cohort | T2D | Non-Disaster (High background) | PCDD/Fs (22.35 pg/g lipid) | Age, survey year, BMI, smoking, and alcohol consumption | OR (95% CI) = 4.98 (1.17-21.17) |
| Female – only studies | Berg et al. 2021 | Norwegian Women and Cancer Study | n=88 | 52 | Case-control | T2D | Non-disaster (low background) | DL-PCBs (21.5 ng/g lipid) | BMI, weight change, breastfeeding, intake of seafood, and total lipids | OR (95% CI) = 2.24 (1.0-5.0) |

| | | | | | | | | | | |
|---|---|---|---|---|---|---|---|---|---|---|
| | Rylander et al. 2015 | Norwegian Women and Cancer Study (NOWAC) cohort) | n=212 | 56 | Case-control | T2D | Non-disaster (low background) | PCB-118 (18.8 ng/g lipid) | BMI, smoking, hypertension, breastfeeding | OR (95% CI) = 1.55 (0.41-5.86) |
| | Zong et al. 2018 | US Nurses' Health Study II (NWSII) Cohort (11-year follow-up) | n=1,586 | 45 | Case-control | T2D | Non-disaster (low background) | DL-PCBs (13 ng/g lipid) | Age, date/time of collection, race/ethnicity, fasting status when blood was drawn, menopausal status, and hormone replacement therapy use | OR (95% CI) = 1.36 (1.01-1.84) |

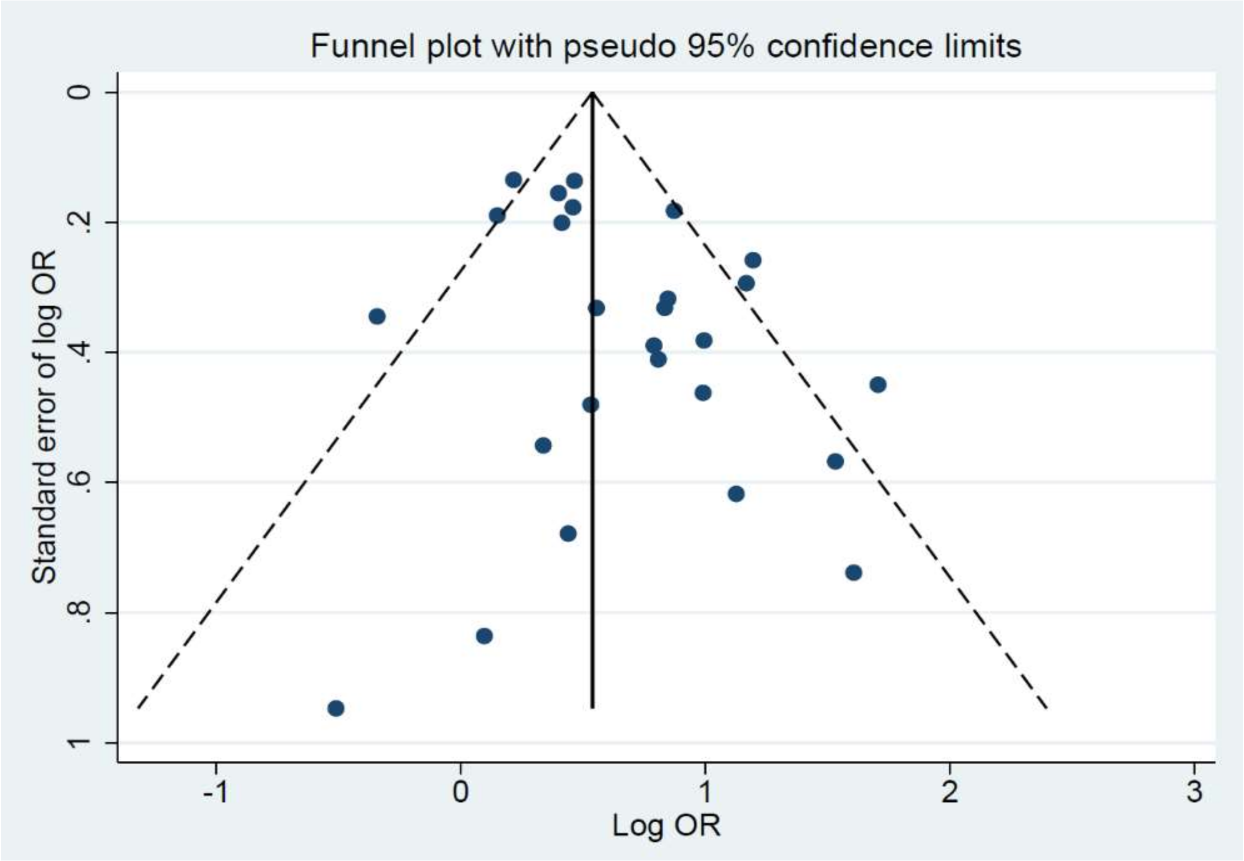


Number of studies = **26**                    Root MSE = **1.356**

| Std_Eff | Coefficient | Std. err. | t | P>\|t\| | [95% conf. | interval] |
|---|---|---|---|---|---|---|
| slope | **.2797132** | **.1423604** | **1.96** | **0.061** | **-.0141042** | **.5735306** |
| bias | **1.124382** | **.5427072** | **2.07** | **0.049** | **.0042893** | **2.244474** |

Test of H0: no small-study effects          P = **0.049**

**Supplementary Figure 1.** Funnel plot (Egger's test) of all studies included in meta-analysis

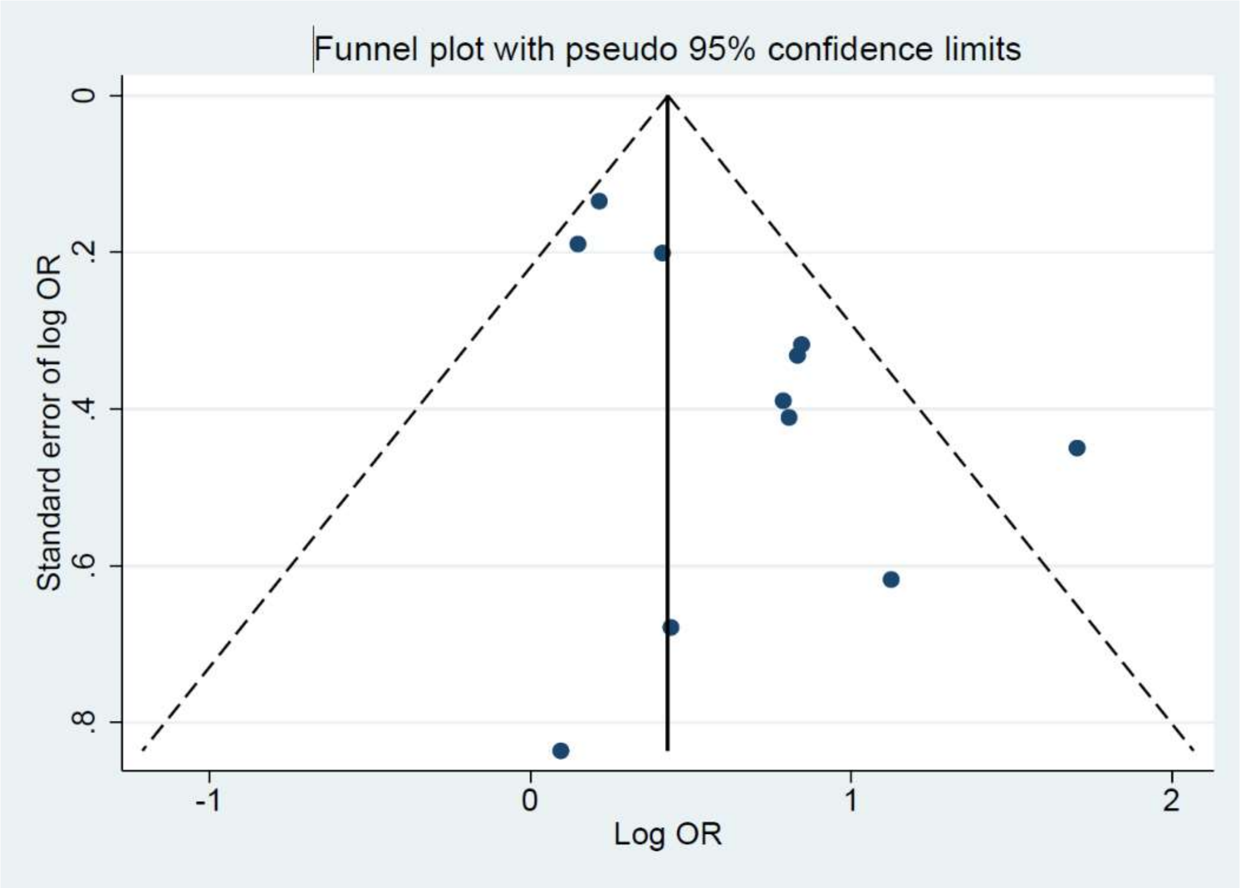


Number of studies = **11** Root MSE = **1.085**

| Std_Eff | Coefficient | Std. err. | t | P>\|t\| | [95% conf. | interval] |
|---|---|---|---|---|---|---|
| slope | **.0116728** | **.1787349** | **0.07** | **0.949** | **-.3926535** | **.4159992** |
| bias | **1.77845** | **.6615225** | **2.69** | **0.025** | **.2819817** | **3.274918** |

Test of H0: no small-study effects P = **0.025**

**Supplementary Figure 2.** Funnel plot (Egger's test) of female-specific data from all studies included in meta-analysis

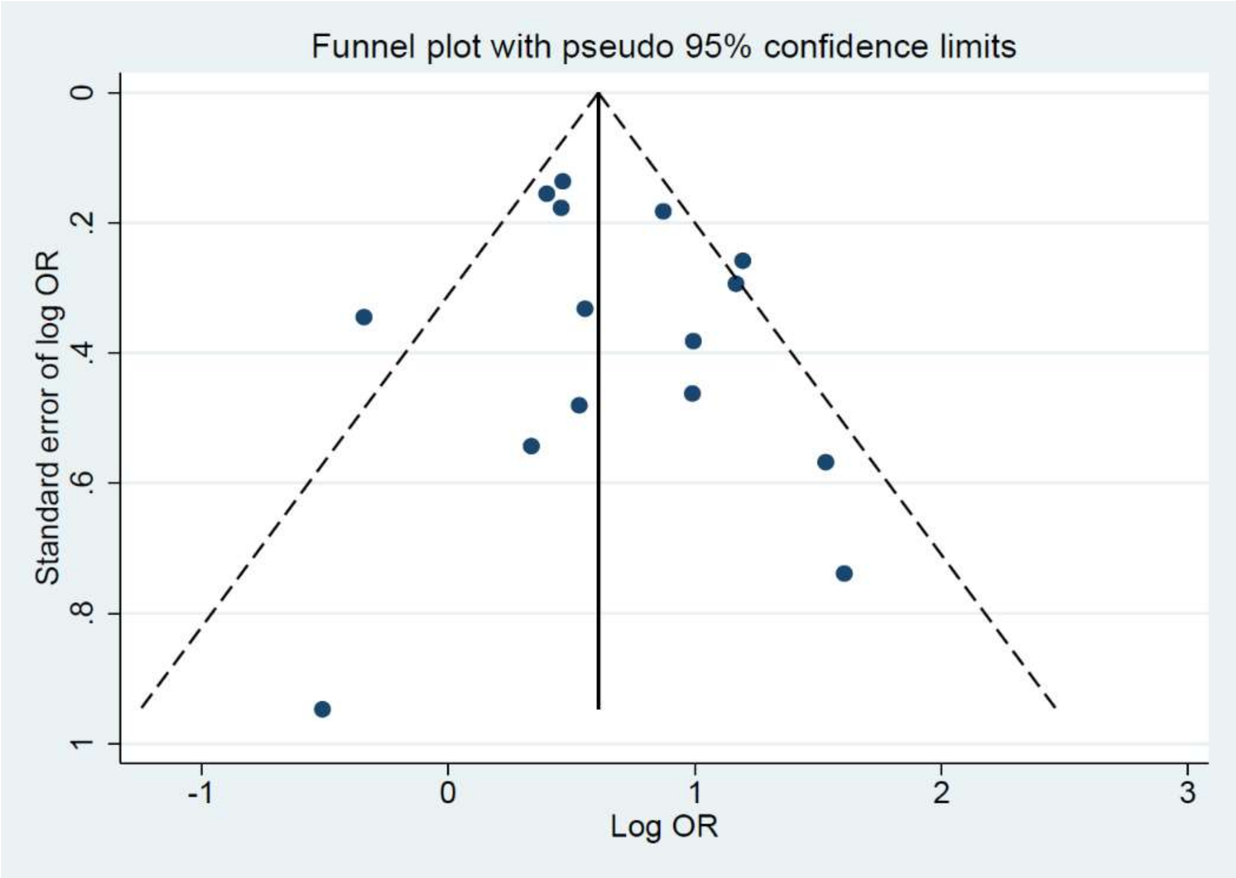


```
Number of studies =  15                          Root MSE      =   1.478

     Std_Eff   Coefficient  Std. err.       t    P>|t|     [95% conf. interval]

       slope     .4561517   .2017619     2.26    0.042     .0202717    .8920318
        bias     .6768291   .7851852     0.86    0.404    -1.01946     2.373119

Test of H0: no small-study effects            P = 0.404
```

**Supplementary Figure 3.** Funnel plot (Egger's test) of male-specific data from all studies included in meta-analysis
